# Sex disparities and neutralizing antibody durability to SARS-CoV-2 infection in convalescent individuals

**DOI:** 10.1101/2021.02.01.21250493

**Authors:** Alena J. Markmann, Natasa Giallourou, D. Ryan Bhowmik, Yixuan J. Hou, Aaron Lerner, David R. Martinez, Lakshmanane Premkumar, Heather Root, David van Duin, Sonia Napravnik, Stephen D. Graham, Quique Guerra, Rajendra Raut, Christos J. Petropoulos, Terri Wrin, Caleb Cornaby, John Schmitz, JoAnn Kuruc, Susan Weiss, Yara Park, Ralph Baric, Aravinda M. de Silva, David M. Margolis, Luther A. Bartelt

## Abstract

The coronavirus disease 2019 (COVID-19) pandemic, caused by severe acute respiratory syndrome-related coronavirus-2 (SARS-CoV-2) has now caused over 2 million deaths worldwide and continues to expand. Currently, much is unknown about functionally neutralizing human antibody responses and durability to SARS-CoV-2. Using convalescent sera collected from 101 COVID-19 recovered individuals 21-212 days after symptom onset with forty-eight additional longitudinal samples, we measured functionality and durability of serum antibodies. We also evaluated associations between individual demographic and clinical parameters with functional neutralizing antibody responses to COVID-19. We found robust antibody durability out to six months, as well as significant positive associations with the magnitude of the neutralizing antibody response and male sex. We also show that SARS-CoV-2 convalescent neutralizing antibodies are higher in individuals with cardio-metabolic comorbidities.

**Significance:** In this study we found that neutralizing antibody responses in COVID-19 convalescent individuals vary in magnitude but are durable and correlate well with RBD Ig binding antibody levels compared to other SARS-CoV-2 antigen responses. In our cohort, higher neutralizing antibody titers are independently and significantly associated with male sex compared to female sex. We also show for the first time, that higher convalescent antibody titers in male donors are associated with increased age and symptom grade. Furthermore, cardio-metabolic co-morbidities are associated with higher antibody titers independently of sex. Here, we present an in-depth evaluation of serologic, demographic, and clinical correlates of functional antibody responses and durability to SARS-CoV-2.

## Introduction

Over twelve months have passed since the emergence and eventual global spread of the novel coronavirus, SARS-CoV-2, the agent of the COVID-19 pandemic. As SARS-CoV-2 continues to spread and mutate across naïve and previously exposed populations, increased understanding of the breadth and durability of individual humoral responses to natural infection is needed to assess the re-infection risk of individuals and also to guide the deployment and to inform recently authorized vaccines and antibody-based therapies. Recent work has shown that SARS-CoV-2 can stimulate the production of highly neutralizing antibodies directed against the spike protein (S) which is necessary for viral attachment, fusion and entry into host cells(1, 2). We and others have shown that antibodies directed against the ACE2 receptor binding domain (RBD) of the S protein consistently demonstrate a strong correlation with functional neutralization (3-5), and are protective in non-human primate and rodent models (6-9). Furthermore, low conservation between the RBD of SARS-CoV-2 and other non-SARS human betacoronaviruses, makes RBD an appealing target for highly specific COVID-19 responses.

Serum antibody responses to endemic betacoronaviruses initially wane weeks to months after infection, but remain detectable up to at least 1 year (10, 11). After SARS-CoV-1 and MERS infections, IgG levels peak at four months, then slowly wane but remain detectable for at least two years and up to 17 years (11, 12). Although antibody seroconversion to primary SARS-CoV-2 infection is nearly universal within the first two weeks after symptom onset (4, 13-15), the magnitude of this response varies with symptom severity (4, 16, 17). Longevity of serum antibodies to SARS-CoV-2 S protein after vaccination as well as natural infection has been studied out to three months, during which time IgG, IgM and IgA levels to most SARS-CoV-2 antigens peak and begin to decline (16, 18-20), as plasmablast and short-lived plasma cell responses wane. More recent data suggests that S protein IgG levels begin to reach a steady level with much slower rates of decline after 90 days post infection (5, 21, 22). Few studies have described long-term durability of SARS-CoV-2 wild-type (WT) virus neutralizing antibodies in recovered individuals, and the protective titer of these antibodies is unknown.

The clinical and demographic determinants of the breadth and durability of functionally neutralizing antibodies, have not been studied in-depth after SARS-CoV-2 infection. A recent study found higher ratios of RBD antibodies to nucleocapsid (N) antibodies in outpatient compared to inpatient populations (4). Another study found positive correlations with RBD and neutralizing antibody levels with male sex, age and symptom severity in a mild disease cohort 60 days post symptom onset (23). Finally, studies have suggested that there is a faster decline in S antibody levels acutely after infection in asymptomatic individuals, compared to symptomatic individuals (4, 17). These early findings in binding and neutralizing antibody levels within the first 30-60 days post infection, indicate that there are significant demographic and clinical differences in humoral immune responses to COVID-19 infection. Identifying these differences is critical to understanding long term protection from natural infection as well as vaccine-induced immunity. In this study, we use both novel and established assays to characterize the binding and longevity of serum antibodies to SARS-CoV-2 RBD, spike protein N-terminal domain (NTD) and N antigens, and to measure the level and durability of SARS-CoV-2 neutralizing antibodies. We further define demographic and clinical correlates of the magnitude and durability of both binding and functional antibody responses to SARS-CoV-2.

## Results

### Donor characteristics

Between April 11^th^ and July 22^nd^ 2020, a total of 101 eligible COVID-19 convalescent plasma (CP) donors were enrolled in this study. The majority of donors donated once, however 31 donors provided sequential donations amounting in an additional 48 serum samples. Donors were over 18 years of age, 51% male and 49% female (based on sex assigned at birth). The median age was 43 years (Interquartile Range 29, full range 18-79), which is similar to other CP donor cohorts (22, 23) and the majority identified as non-Hispanic, white/Caucasian. Donors were diagnosed with COVID-19 by either SARS-CoV-2 reverse-transcriptase polymerase chain reaction (RT-PCR) (*n* = 79) or blood antibody testing by EUA approved commercial assays (*n* = 22) (Table 1 & Supplementary Table 1). Donors diagnosed by antibody test had either RT-PCR-confirmed household contacts, COVID-19 symptoms without RT-PCR testing, or unable to provide a copy of their RT-PCR result. The median time from symptom onset or RT-PCR diagnosis to first donation was 57 days (full range 21-121). Thirty-four donors reported comorbid conditions, the most common being hay fever and high blood pressure (Supplementary Table 2). Eight donors were asymptomatic and 93 reported symptoms. The median time of symptom duration for symptomatic donors without ongoing symptoms (72/90) was 16 days (full range 2-107). Fifty-seven donors had mild-to-moderate disease (Grade 1-2; outpatient), 14 donors had severe disease (Grade 3-4; hospitalized), and 22 donors had unknown disease severity (Table 1). The most common symptoms reported were fatigue (89%), headache (77%), fever (74%) and muscle aches (73%) (Supplementary Table 3). The majority of donors resided in central North Carolina, with the highest proportion from Orange and Wake counties (Supplementary Fig. 1).

**Table 1.**
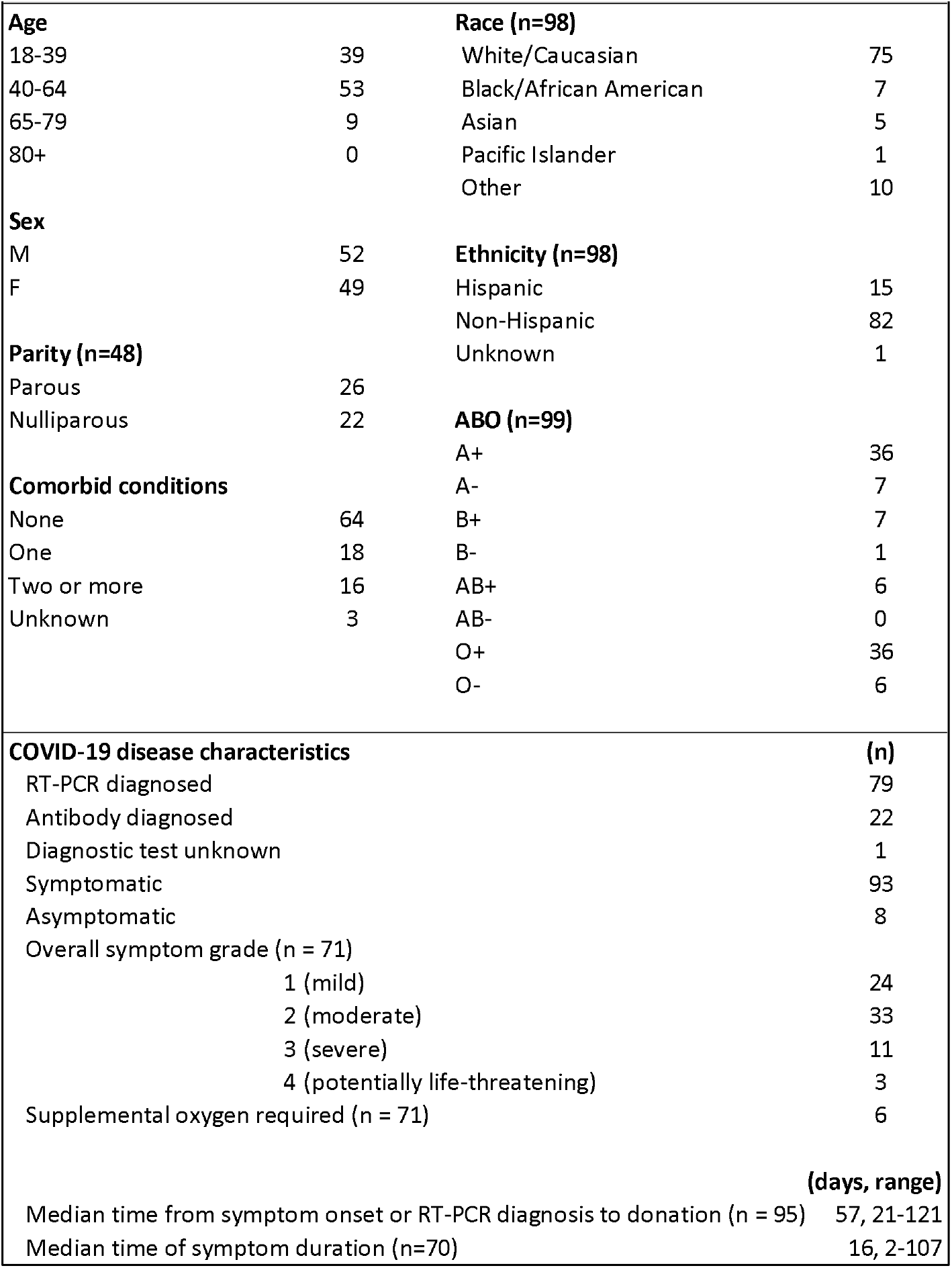
Convalescent plasma donor characteristics at time of donation (n=101 unless otherwise specified). Plasma donor demographic and COVID-19 disease characteristics. RT-PCR; Reverse-transcriptase polymerase chain reaction. Symptom grades: GRADE 1 MILD; Mild symptoms causing no or minimal interference with usual social & functional activities with intervention not indicated. GRADE 2 MODERATE; Moderate symptoms causing greater than minimal interference with usual social & functional activities with intervention indicated. GRADE 3 SEVERE; Severe symptoms causing inability to perform usual social & functional activities with intervention or hospitalization indicated. Oxygen administered via nasal cannula. GRADE 4 POTENTIALLY LIFE-THREATENING; potentially life-threatening symptoms causing inability to perform basic self-care functions with intervention indicated to prevent permanent impairment, persistent disability, or death. Hospitalization requiring intubation or use of supplemental oxygen (CPAP or oxygen administered via mask).

| Table 1. Convalescent plasma donor characteristics at time of donation<br>(n=101 unless otherwise specified) |  |  |  |
| --- | --- | --- | --- |
| **Age** |  | **Race (n=98)** |  |
| 18-39 | 39 | White/Caucasian | 75 |
| 40-64 | 53 | Black/African American | 7 |
| 65-79 | 9 | Asian | 5 |
| 80+ | 0 | Pacific Islander | 1 |
|  |  | Other | 10 |
| **Sex** |  | **Ethnicity (n=98)** |  |
| M | 52 | Hispanic | 15 |
| F | 49 | Non-Hispanic | 82 |
| **Parity (n=48)** |  | Unknown | 1 |
| Parous | 26 | **ABO (n=99)** |  |
| Nulliparous | 22 | A+ | 36 |
| **Comorbid conditions** |  | A- | 7 |
| None | 64 | B+ | 7 |
| One | 18 | B- | 1 |
| Two or more | 16 | AB+ | 6 |
| Unknown | 3 | AB- | 0 |
|  |  | O+ | 36 |
|  |  | O- | 6 |
| **COVID-19 disease characteristics** |  |  | **(n)** |
| RT-PCR diagnosed |  |  | 79 |
| Antibody diagnosed |  |  | 22 |
| Diagnostic test unknown |  |  | 1 |
| Symptomatic |  |  | 93 |
| Asymptomatic |  |  | 8 |
| Overall symptom grade (n = 71) |  |  |  |
| 1 (mild) |  |  | 24 |
| 2 (moderate) |  |  | 33 |
| 3 (severe) |  |  | 11 |
| 4 (potentially life-threatening) |  |  | 3 |
| Supplemental oxygen required (n = 71) |  |  | 6 |
|  |  |  | **(days, range)** |
| Median time from symptom onset or RT-PCR diagnosis to donation (n = 95) |  |  | 57, 21-121 |
| Median time of symptom duration (n=70) |  |  | 16, 2-107 |

### Neutralization and binding antibody assays

To investigate in-depth functional antibody responses to SARS-CoV-2 infection at convalescence, we employed two virus neutralization assays, one using an authentic WT SARS-CoV-2 with a luciferase reporter(24), and another using a PSV assay (see Methods). We also measured total Ig binding to the spike protein RBD and NTD, as well as IgG binding to N protein antigen. We found that 98% (99/101) of donors generated antibodies to at least one SARS-CoV-2 antigen or virus (Fig. 1a,b), 92% (93/101) had at least two positive antibody assays, and 65% (65/101) had functional and binding antibodies to all viruses and antigens. Only two donors had negative results in every assay, both were asymptomatic and both diagnosed by an antibody test. We found that the most sensitive assays to detect antibodies in recovered donors were the RBD total Ig assay (96% of donors positive), followed by PSV neutralization and N IgG assays (both 93% of donors positive).

**Fig. 1.**
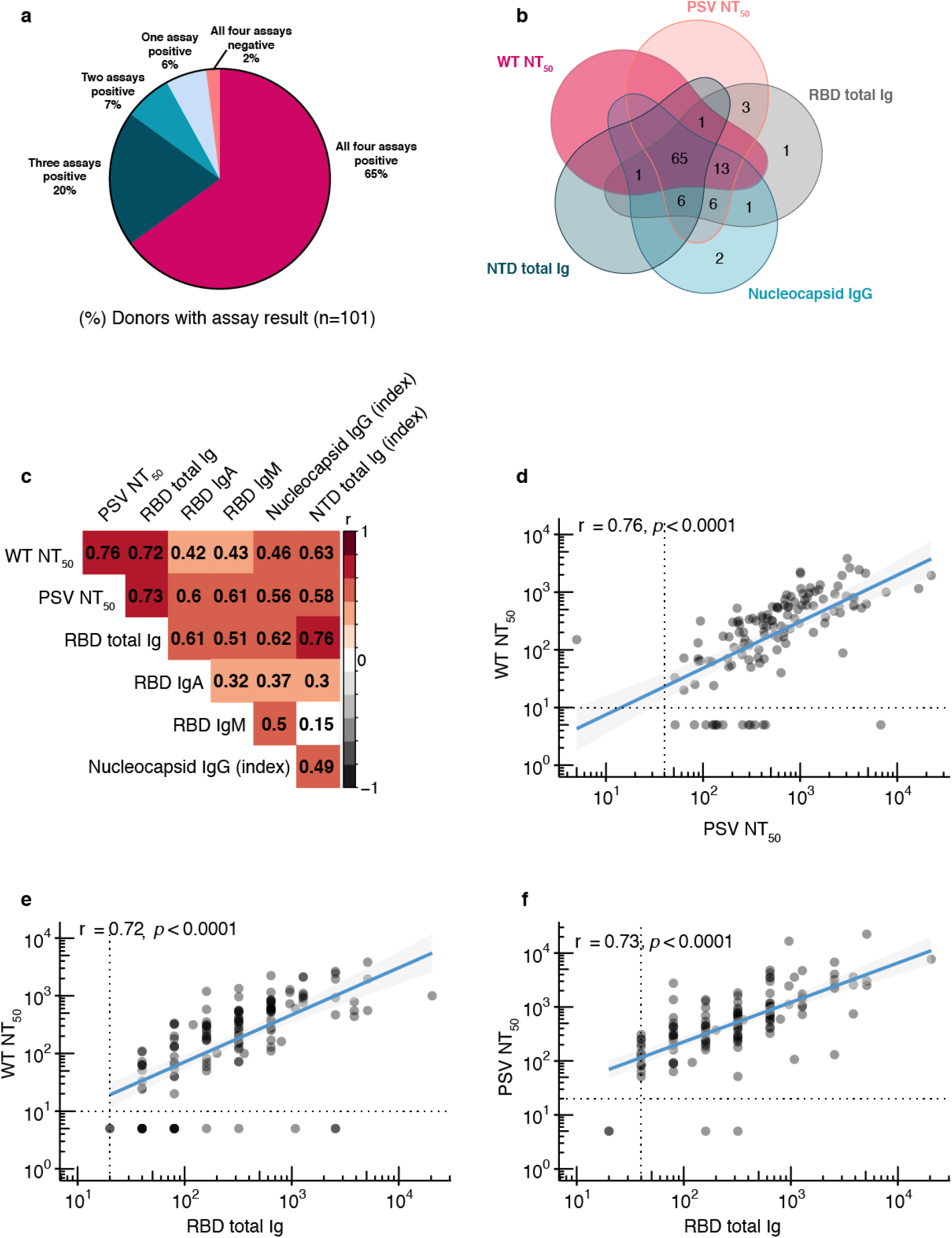
Neutralizing and binding antibody results. **a**, Pie chart with overall assay results for all 101 donors, four assays shown (wild-type neutralization assay, RBD and NTD total Ig assays, Nucleocapsid IgG assay), **b**, Venn diagram showing overlap among five assays (wild-type neutralization assay, pseudovirus neutralization assay, RBD and NTD total Ig assays, Nucleocapsid IgG assay), **c**, Heat map of Spearman’s correlation coefficients examining the association between all assays performed. Red colour represents positive association between assays and black represents negative associations. Not significant correlations coefficients (p>0.05) are left blank, **d**, Wild-type virus NT50 dilution plotted against pseudovirus NT50 dilution, p<0.0001, **e**, Wild-type virus NT50 dilution plotted against RBD total Ig antibody level (end-point titer), p<0.0001. **f**, Pseudovirus NT50 dilution plotted against RBD total Ig antibody level (end-point titer), p<0.0001. For **d-f**, non-parametric, two-tailed Spearman’s rank correlation was used to calculate correlation coefficients (r) and P values (p), titers below LOD set to 5, all double-negative values removed, blue lines represent linear regression fit with 95% confidence interval (gray shading).

All donors with undetectable RBD antibody titers also had undetectable neutralizing antibody assays, and the RBD binding assay showed the strongest correlation with the two neutralization assays (Fig. 1c-f, Supplementary Fig. 2). Among the other binding assays, the N assay had the weakest correlations with both neutralization assays compared to the spike antigen based NTD assay. We then looked at quantitative measures of functionally neutralizing as well as RBD-binding antibody levels by end-point titer. The majority of donors (80%) had detectable WT neutralizing antibody titers, and > 50% of these exceeded 1:160 (the FDA-recommended threshold for therapeutic applications of convalescent plasma) (Supplementary Fig. 3). The majority of RBD total Ig end-point titers were found to be within the range of 1:160-1:640. Since isotype specific IgA and to a lesser extent, IgM antibodies may influence the early neutralizing antibody response (25), we also measured RBD IgA and IgM binding titers. Approximately 60% of donors demonstrated detectable IgA or IgM antibodies to RBD, with most in the lower titer range (1:20-1:159) (Supplementary Fig. 3).

### Functional and binding antibody level durability

Overall donor antibody levels, including additional donations from 31/101 donors who donated more than once (total samples donated n=149), revealed stable neutralizing, RBD, and NTD-binding antibodies over six months (Fig. 2). Among the specific assays, neutralizing antibodies to WT virus and binding antibodies to NTD were the most stable out to 180 days (Fig. 2a,b). Through 120 days and beyond, there was a slight decrease in PSV neutralizing antibodies and total Ig binding antibodies to RBD (Fig. 2c,d). RBD total Ig decreases over time were likely due in part to the decline of IgA and IgM titers that we observed after day 90 (Fig. 2e,f). Notably, compared with antibodies directed against spike protein antigens, there was a stronger decrease in N binding IgG levels over this time period (Fig. 2g). When comparing the correlation coefficients of the trendlines in Fig. 2a-d,g with a Fisher r-to-z transformation, we found significant (p < 0.05) differences only between NTD antibody levels compared to all of the other assays which either stay constant or decrease.

**Fig. 2.**
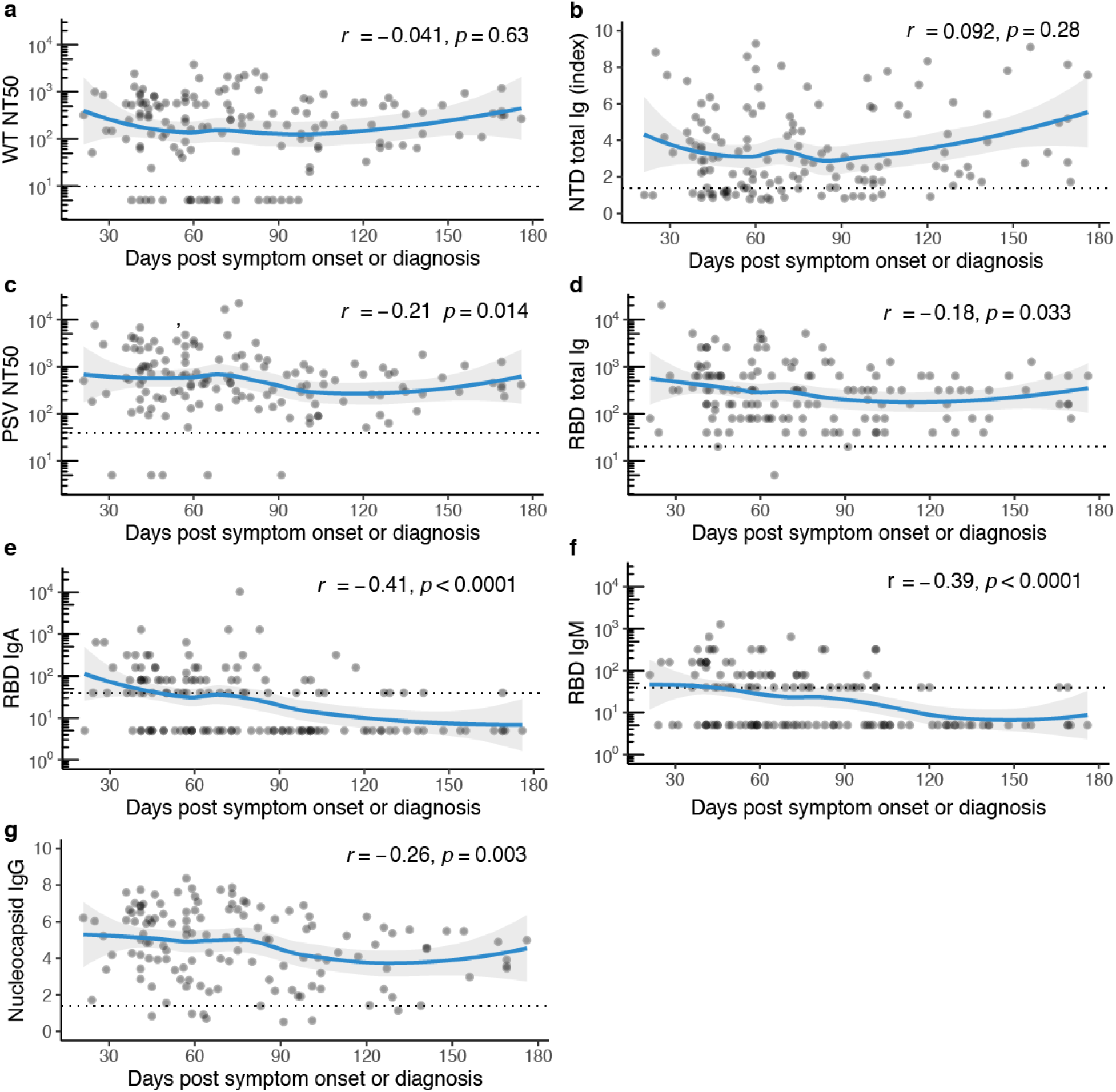
Antibody titers over time. **a**, Functional antibody (WT NT50 dilution) plotted against days post symptom onset or RT-PCR diagnosis, r=-0.041, p = 0.63, **b**, Functional antibody (PSV NT50 dilution) plotted against days post symptom onset or RT-PCR diagnosis, r=-0.21, p=0.014. **c**, Nucleocapsid IgG (index value) plotted against days post symptom onset or RT-PCR diagnosis, r=0.092, p=0.0029, **d**, NTD total Ig (P/N ratio) plotted against days post symptom onset or RT-PCR diagnosis, r=0.092, p=0.28, **e**, RBD total Ig (end-point titer) plotted against days post symptom onset or RT-PCR diagnosis, r=-0.18, p=0.033, **f**, RBD IgM (end-point titer) plotted against days post symptom onset or RT-PCR diagnosis, r=-0.39, p<0.0001, **g**, RBD IgA (end-point titer) plotted against days post symptom onset or RT-PCR diagnosis, r=-0.41, p<0.0001. For **a-g**, n=138, non-parametric, two-tailed Spearman’s rank correlation was used to calculate correlation coefficients (r) and P values (p), titers below LOD set to 5, all double-negative values removed, blue lines represent loess regression fit with 95% confidence interval (gray shading).

We then studied in detail the donors who provided sequential donations to examine temporal kinetics of antibody levels at an individual level. Overall functional neutralizing antibody levels to WT virus and RBD-binding Ig levels showed no significant changes between donation times (Fig. 3a,b and Supplementary Fig. 4). To ascertain if initial antibody titer plays a role in antibody changes over time, we separated sequential donors into three groups by initial titer: > 1:640, 1:160-1:640, and 1:20-1:159. Median WT viral neutralization antibody titers (Fig. 3c-e) and RDB Ig antibody titers (Fig. 3f-h) between the first two donations showed a modest decrease in the highest initial titer group (>1:640), but not in the lower titer groups. However, earlier time points are needed in the lower titer groups to better compare these levels to the high titer group, as we may not see changes in the lower titer groups due to longer time to first donation in these groups. This decrease in the RBD total Ig group with initial titer > 1:640 was likely due to RBD IgA and IgM levels in these donors, which showed a significant decline between the first two donations (p < 0.05) (data not shown).

**Fig. 3.**
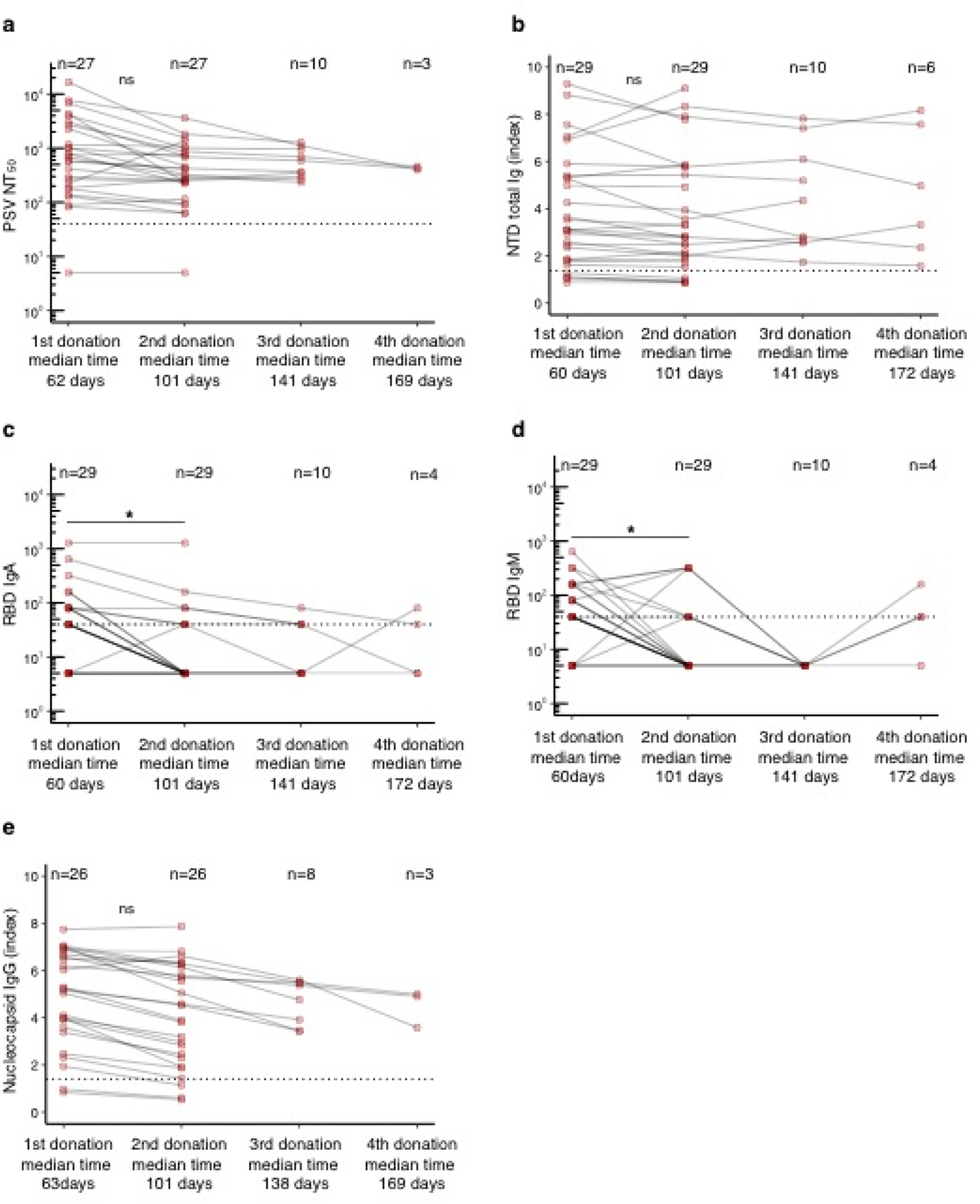
Antibody titers over time in sequential donors. **a**, Functional antibody (NT50 dilution) of sequential donors over four donations. **b**, RBD total Ig titers of sequential donors over four donations. **c-e**, Functional antibody (NT50 dilution) stratified by titer levels at first donation. **f-h**, RBD total Ig stratified by titer levels at first donation. Titers are presented as geometric mean with geometric coefficient of variation. Statistical significance was determined using non-parametric Kruskal-Wallis test adjusted for multiple comparisons for **a-b**. For **c-h** statistical significance was determined using Mann-Whitney U-tests comparing donation 1 vs donation 2 for which matching donor data was available.

### Demographic and clinical correlates of functional antibody titers

SARS-CoV-2 binding and functionally neutralizing antibody levels were higher in males compared to females (Fig. 4a,b, Supplementary Fig. 5), and increased with increasing age and correlate positively with male age and symptom grade (Fig. 4c,d). This difference between males and females (Fig. 4a,b) remained after negative data points were removed from each analysis. Surprisingly, positive correlations with antibody levels and age and symptom grade (Fig. 4c,d, Extended Data Fig. 4) were restricted to the male population (Fig. 4e,f). Sex stratification revealed that in males, age and symptom grade were significantly positively correlated, as were age and RBD Ig and functionally neutralizing antibody levels (Fig. 4e,g-k). On the other hand, in females, only RBD IgA levels were associated with symptom grade (Fig. 4f). Males and females were equally likely to be hospitalized (p = 0.95).

**Fig. 4.**
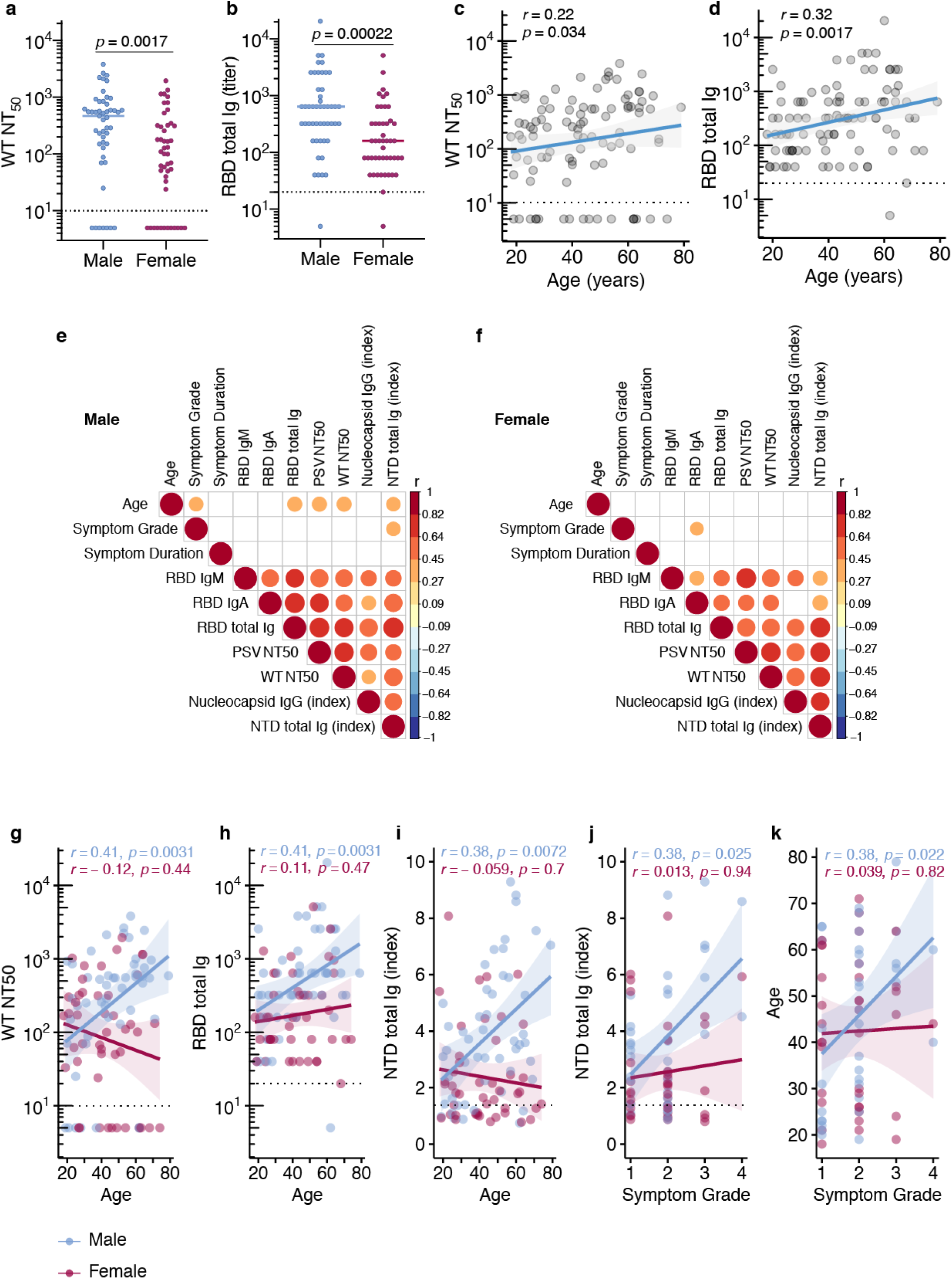
Clinical correlates of antibody titers. **a**, Functional antibody (NT50 dilution) in males (n=49) and females (n=46) at first donation, **b**, RBD total Ig titers in males and females at first donation. Horizontal bars indicate median values. For **a**,**b** statistical significance was determined using Mann-Whitney U-tests, **c**,**d**, Spearman’s correlation between age correlation between age and NT50 or RBD Ig levels at first donation, **e**,**f**, non-parametric, two-tailed, Spearman’s correlations heat map of clinical correlates and antibody titers stratified by sex, (red = positive association, blue = negative association, blank = not significant association), **g-k**, correlation between age and NT50 or RBD Ig levels at first donation. Spearman’s rank correlation was used to calculate correlation coefficients (r) and P values (p)

We then examined the possibility that antibody stability over time was influenced by sex or age. No significant differences were observed in WT neutralizing antibody levels or RBD Ig levels over time (first 90 days) between males and females (Fig. 5a,b). In contrast, there were rapid declines in both types of antibodies in the youngest age group (18-43yo) over the first 90-day period (Fig. 5c,d) that may have been related to decreases in serum RBD IgA, but not IgM, which showed a significant decline in this age group over this time period (Fig. 5e). We then calculated an estimate of the effect of age, adjusted for time from symptom onset to donation, stratified by sex on the various functional and binding antibody levels. Among males we observed that for each one year increase in age there was a significant increase in antibody levels in all assays tested except N IgG and RBD IgM, but among females age did not seem to affect antibody levels after accounting for time from symptom onset (Fig. 5f).

**Fig. 5.**
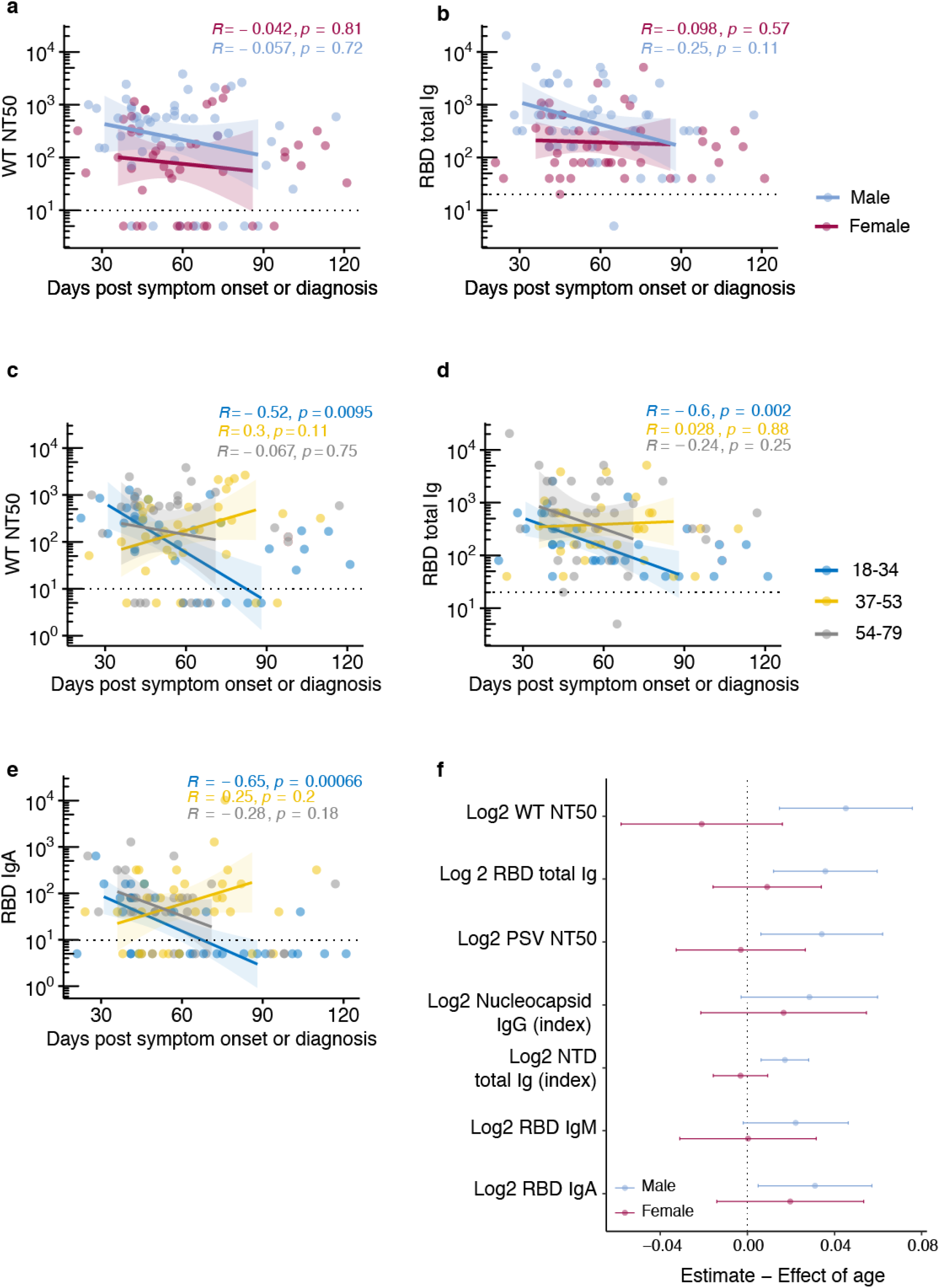
Antibody differences between sexes and age groups. **a**, Differences in functional antibody (NT50 dilution) levels between males (n=49) and females (n=46) at first donation. **b**, Differences in RBD total Ig titers between males and females at first donation. **c**, Differences in functional antibody (NT50 dilution) levels between age groups. **d**, Differences in in RBD total Ig titers between age groups. For **c-d** Donors were divided into tertiles based on their age. For **a-d**, lines represent linear regression fit and shaded areas represent 95% confidence interval. Lines from linear regression were fitted from day 30-90 to avoid overfitting where fewer observations were available. Spearman’s rank correlation was used to calculate correlation coefficients (r) and P values (p) e, Forest plot of estimated effect (95% CI) of age on antibody titers at first donation, stratified by sex. Linear regression model was adjusted for time from symptom onset or RT-PCR diagnosis.

Since we identified that in male donors, increased symptom grade, or disease severity correlated with higher antibody levels, we looked more closely at individual symptoms to ascertain if any in particular were associated with each other, or with donor serum antibody levels. We found that of the most common symptoms, only loss of sense of taste and smell were associated, though more strongly in female than male donors (Supplementary Fig. 7a, b). Surprisingly, we found a negative effect of reporting tiredness or fatigue in male donors on the level of RBD Ig binding antibodies (Supplementary Fig. 7b). We also evaluated the association between antibody levels and the presence of comorbid conditions and found that donors with cardio-metabolic diseases had higher levels of neutralizing, RBD Ig, N IgG and NTD Ig antibodies. This observation was independent of sex. Symptom duration (Supplementary Fig. 5k-q,4), nulliparity and ABO blood group were not significantly associated with functional or binding antibody levels.

## Discussion

In-depth serological, clinical and demographic correlates of durable and protective functional antibodies in individuals who have recovered from COVID-19 have not been well described. Understanding serological responses to COVID-19 disease and vaccination, will allow us to define which antibody populations may be protective against reinfection and thus act as immunological correlates of protection. Of 101 convalescent plasma donors who experienced a range of COVID-19 disease, the vast majority have detectable levels of functionally neutralizing as well as binding antibodies to SARS-CoV-2 RBD, NTD and N antigens. Furthermore, though their titers are heterogeneous, most donors have neutralizing and RBD-targeting antibody titers of >1:160. Even low levels of functionally neutralizing antibodies to SARS-CoV-2, as seen here in about a quarter of donors, are protective in non-human primate vaccine models (26, 27). This suggests that low serum levels of a few highly potent antibodies may be enough to confer protection, and we find that such antibodies are nearly universally produced upon exposure to the virus in this donor cohort of mostly symptomatic cases.

Of three SARS-CoV-2 antigens used for antibody detection in this study, the RBD was the most sensitive in detecting prior SARS-CoV-2 infection. Furthermore, RBD total Ig levels showed the strongest correlation with functionally neutralizing antibodies, suggesting its role as the immunodominant antigenic target of antibodies that neutralize SARS-CoV-2 infection. We found that 95% of sera with an RBD total Ig titer of ≥1:160 had positive WT virus neutralizing antibody titers, suggesting that this may be a cutoff used as a surrogate for functional antibody assays. Furthermore, the majority of donors with undetectable WT virus neutralizing antibody levels, had detectable RBD-binding antibodies, suggesting they may have RBD-targeting neutralizing antibodies that are below the assay detection limit. This hypothesis can be tested in future studies using passive transfer mouse protection models. This highlights the potential role for RBD-based antibody assay development and testing as a surrogate for functional antibody assays that could be deployed in the clinical and vaccination setting in a scalable, high-throughput fashion.

The strongest demographic correlate of neutralizing antibody levels we found was male sex. Studies have shown that COVID-19 disease is associated with higher morbidity and mortality rates in men compared with women (28). The reason for this finding is unknown, and seems unrelated to CD8+ and CD4+ T cell frequency (22). Sex differences in other respiratory viral disease outbreaks have been seen, for example during the 2009 influenza pandemic where female sex correlated with severe disease in a young cohort in Canada(29). Some viral infections as well as vaccinations such as the influenza vaccination have been seen to elicit stronger serum antibody and cellular immune responses in females(30), while others elicit stronger serologic antibody responses in males(31). Differences in disease severity and humoral responses to vaccines have been hypothesized to be influenced by a combination of sex hormone effects on immune cell signaling, X chromosome immune-related gene expression and miRNA levels, and genetic polymorphisms (30) in genes encoding important immunologic proteins such as IL-6 (32) and CTLA-4 (33).

Robbiani *et al*. reported significantly higher SARS-CoV-2 RBD antibodies and functionally neutralizing antibodies in male compared to female COVID-19 CP donors with mild-moderate disease in the first 60 days post symptom onset (23). Similarly, we find significant correlations of higher functional and binding antibody levels with male sex, continued out to 180 days. We further add that these sex differences in antibody levels are also seen with SARS-CoV-2 N protein and NTD antigens, and that age and symptom grade also influenced the sex disparity in RBD binding and functionally neutralizing antibody responses. These findings suggest that male sex, especially males with increased age and worse COVID-19 symptom severity may be a demographic and clinical correlate of functional antibodies. Studies have shown that early convalescent functional antibody levels are higher in individuals with more severe COVID-19(16, 19), which may be a product of prolonged viral replication and immune antigenic exposure. Our findings recognize that there are currently unknown underlying factors which predispose older males to either prolonged viral replication and immune exposure to SARS-CoV-2, or differential immune activation.

We do not yet know what level of functional antibodies is required for protection from SARS-CoV-2. Although female donors in this cohort have lower antibody levels than males, this may be enough to confer long-term protection. This observation warrants further investigation, including consideration of a similar sex-bias in vaccine-induced immunity. Furthermore, we found significant differences in sex and functional antibody production despite reported disease severity, suggesting that pro-longed viremia and/or abnormal cytokine activation may not be the only things responsible for this finding. Other hypotheses that have been made to explain COVID-19 disease sex differences include poorer T cell responses in males (28), and the presence of previously undetected auto-antibodies against Type I interferons (34) in males with severe disease. On the other hand, the hypothesis that expression of ACE2 and TMPRSS, important SARS-CoV-2 cellular entry receptors in human lung and other tissues, play a role in the sex disparity is thought to be an unlikely explanation (35).

In the face of COVID-19 vaccinations and new viral mutations, it is critical to define functional antibody durability after natural infection and vaccination. Here we show for the first time, that functionally neutralizing antibodies to WT SARS-CoV-2 virus remain stable months post symptom onset, and that this is likely maintained to 180 days. Not surprisingly, levels of RBD-binding IgA and IgM antibodies declined rapidly within the first three months after symptom onset. However, NTD-binding Ig antibodies remain stable, and RBD-binding Ig antibodies declined modestly. Levels of WT neutralizing and RBD Ig antibodies on an individual level were also maintained, with only a modest decrease within the first 90 days after symptom onset in donors with initial titers > 1:640. When broken down by age group, 18-34 year old donors demonstrated a significant decrease in functional antibody and RBD Ig responses over the first 90 days post symptom onset that was likely driven by rapidly declining RBD IgA levels.

We also find that N IgG antibodies correlate least with neutralizing antibodies and continue to decline 120-180 days post symptom onset, a trend which was noted 90 days post symptom-onset in a mild-disease community cohort (18). This suggests that though SARS-CoV-2 N antibodies may be generated at high levels early after symptomatic infection, N may not be an immunodominant target of the adaptive immune response, and thus is a less sensitive measure of remote infection. This further suggests that the use of N protein in seroprevalence studies may bias results towards more recent infections and warrants further investigation in cohorts of mild and asymptomatic COVID-19 disease.

One major limitation of this study is the demographic uniformity of our study population, which limits the generalizability of our findings and highlights the need to do these studies with a more diverse and representative population. Another bias in our donor population is our focus on re-calling donors with higher neutralizing antibody titers to repeat donations. Thus, our “sequential donation” population is biased towards higher titer donors.

Understanding human antibody responses and correlates of neutralizing antibodies to SARS-CoV-2 is critical in the next coming phase of understanding SARS-CoV-2 vaccine efficacy and protection against reinfection. We find that WT SARS-CoV-2 functionally neutralizing antibodies are maintained for months after infection. Our findings further support the role of RBD binding antibodies as correlates of functionally neutralizing antibodies, suggesting that vaccines that induce potent RBD responses may be particularly efficacious. Furthermore, we identify for the first time, a role for sex differences as sustained correlates of WT SARS-CoV-2 functional neutralization. The association of male sex in this cohort with higher neutralizing antibody levels reveals a sexual dimorphism in humoral immune responses to SARS-CoV-2. We hypothesize that this is likely due to a combination of factors such as differences in duration of mucosal replication, T cell responses, sex hormone roles in immune activation, and genetic differences in immune responses. This finding may have clinical as well as vaccine outcome implications, and warrants further investigation.

## Methods

### Donors and Plasma Collection

Convalescent plasma was obtained from volunteer donors who met FDA criteria for plasma collections in the UNC Blood Donor Center. Donors were recruited via IRB-approved direct contact of SARS-CoV-2 positive persons diagnosed through the hospital laboratory system, and public solicitation through multi-media outlets. Fresh sera and plasma collected in the diversion pouch as part of the standard plasmapheresis procedure was saved for research from donors consented to study participation. All donors had confirmed COVID-19 infection by blood antibody testing or nasopharyngeal swab indicating the presence of SARS-CoV-2 RNA as performed by RT-PCR in a US laboratory with a Clinical Laboratory Improvement Amendments certification. All donors were recovered from their COVID-19 illness and qualified for collection in adherence with FDA-regulatory guidance. As required at the time, some donors had a negative repeat SARS-CoV-2 RT-PCR test done within 72 hours prior to donation. At the time of plasma collection, donors were offered participation in the study. All donors who participated provided written informed consent. The research was approved by the UNC Institutional Review Board, and conducted under good clinical research practices. Participating donor characteristics and information regarding COVID-19 symptoms and history were obtained through in-person and telephone interviews using a standardized questionnaire as part of UNC IRB #20-1141. We generated a 4-point symptom severity scale for this study based on the DAIDS grading system(36). For this study time period we did not pre-screen donors to determine presence of SARS-CoV-2 antibodies, donor qualifications were based strictly on their positive COVID-19 diagnostic test and eligibility for plasma donation.

### Recombinant SARS-CoV-2 spike protein antigens

The production of RBD antigen from SARS-CoV-2 was previously described (3). The NTD antigen (16–305 amino acids, Accession: P0DTC2.1) was cloned into the pαH mammalian expression vector and purified using nickel-nitrilotriacetic acid agarose in the same manner.

### Enzyme-linked Immunosorbent Assays

The RBD ELISA assay used in this study was initially described here (3), and the NTD ELISA was performed in the same manner. Briefly ELISAs were done either as a single-point dilution at 1:40 or as serial titrations starting at a dilution of 1:20 or 1:40. ELISA plates were coated with 200ng/well of antigen and blocked, a 2-fold serum dilution series was done and diluted sera was incubated for 1hr at 37°C. Alkaline-phosphatase linked secondary antibodies were used at 1:2500 dilution (IgM and IgG, Sigma; IgA, Abcam). PNPP substrate (Sigma) was added to develop the plate and absorbance was measured at 10 minutes for total Ig, IgG or 25 minutes for IgA or IgM at 405nm using a plate reader (BioTek). Each sample was performed in duplicate. Antibody titration measurements were recorded as end-point titers. Ten plasma samples were tested in the RBD total Ig format and compared to serum, all titer results were within a 2-fold dilution (data not shown). ROC analyses were done to obtain cutoff values and sensitivity and specificity estimates on the SARS-CoV-2 assays using pre-2019 negative control sera and RT-PCR confirmed COVID-19 cases that were at least nine days post-symptom onset (Supplementary Table 4). Positive and negative controls were used to standardize each ELISA assay and normalize across experiments.

### Nucleocapsid protein ELISA

Detection of IgG antibody to SARS-CoV-2 N antigen was performed with a microparticle chemiluminescence assay (Abbott Laboratories) on the Abbott Architect i2000SR immunoassay analyzer. The EUA approved Abbott SARS-CoV-2 IgG assay utilizes microparticles coated with SARS-CoV-2 N protein to capture N specific IgG. Bound IgG was detected via addition of anti-human acridinium-labeled second-step antibody. Following a second wash step, pre-trigger and trigger solutions were added and a chemiluminescent reaction was detected and reported in relative light units (RLU). The RLU generated is reflective of the amount of antibody bound to the microparticles. The sample RLU was compared to the assay-specific calibrator RLU to generate an index value (S/C). Index values >/= 1.4 were considered positive. Sensitivity and specificity has been previously obtained for this assay (Supplementary Table 4) (37, 38).

### SARS-CoV-2-WA1 neutralization assay

Full-length SARS-CoV-2 viruses expressing a nano-luciferase gene were designed and recovered via reverse genetics as previously described (3, 24) in a 96-well micro neutralization format. Briefly, Vero E6 cells were infected with SARS-2-pLuc viruses and titered to generate an 8-point curve. Initial serum dilutions to detect the presence of neutralizing antibody were 1:20 or 1:50, and all serum samples were tested in duplicate. Internal serum controls, cell-only controls, and virus-only controls were included in each neutralization assay plate. Plates were incubated for 48 hours, at which point cells were lysed and luciferase activity was measured on a Nano-Glo Luciferase Assay System (Promega). Antibody neutralization titers to SARS-CoV-2 were reported as serum dilutions at which a 50% reduction in relative light units (NT50) to virus-only controls were observed. LOD was set to 1:10, or ½ the starting dilution of 1:20, since all NT50 values above a titer of 1:10 that were run with a 1:50 starting dilution were > 1:25. Thirteen plasma samples were tested and compared to serum, all NT50 results were within a 3-fold dilution (data not shown). Pre-COVID-19 serum samples were also tested, and 13/13 had NT50 < 1:20 in this assay.

### SARS-CoV-2 Pseudovirus neutralization assay

The “PhenoSense SARS CoV-2 nAb Assay” has been developed by leveraging the proprietary PhenoSense Assay platform that was developed to evaluate antiretroviral drug susceptibility(39) and later adapted to evaluate entry inhibitors and neutralizing antibody(40) as well as co-receptor tropism(41). The production of luciferase is dependent on virus entry and the completion of a single round of virus replication. Agents that inhibit pseudovirus entry or replication reduce luciferase activity in a dose-dependent manner, providing a quantitative measure of drug and antibody susceptibility.

The measurement of neutralizing antibody activity using the PhenoSense SARS CoV-2 nAb Assay is performed by generating HIV-1 pseudovirions that contain and express the complete SARS CoV-2 spike protein open reading frame. The pseudovirus is prepared by co-transfecting HEK293 producer cells with an HIV-1 genomic vector and a SARS CoV-2 envelope expression vector. Neutralizing antibody activity is measured by assessing the inhibition of luciferase activity in HEK293 target cells expressing the ACE2 receptor following pre-incubation of the pseudovirions with serial dilutions of the serum specimen. The expression of luciferase activity in target cells is inhibited in the presence of anti-SARS CoV-2 neutralizing antibody. Data are displayed by plotting the percent inhibition of luciferase activity against log_10_ reciprocal of the serum/plasma dilution. Neutralizing antibody titers are reported as the reciprocal of the serum dilution conferring 50% inhibition (NT50) of pseudovirus infection.

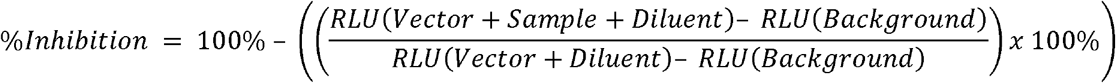

The results of the PhenoSense SARS CoV-2 nAb Assay can be reported as an NT50 titer (1/Dilution) or qualitatively (positive, negative) based on a pre-defined dilution cutoff (e.g. >50% inhibition at 1:40 dilution). To insure that the measured neutralizing antibody activity is SARS CoV-2 specific, each test specimen is also assessed using a non-specific pseudovirus (specificity control) that expresses a non-reactive envelope protein of one or more unrelated viruses (e.g. avian influenza virus).

### Statistical analyses

We used the Wilcoxon rank-sum test to test for differences between two groups and the Kruskal-Wallis test followed by Benjamini-Yekutieli correction to test for differences between three or more groups. We calculated the phi coefficient as a measure of association between two binary factors and relied on the Chi-square test to test for differences. We also calculated the Spearman’s rank correlation coefficient, and used locally estimated scatterplot smoothing (LOESS) to visualize antibody trends over time. Linear regression models were used to further assess relationships with antibody levels, after first transforming antibody levels to the base-2 logarithm scale. Venn diagram and correlation heat maps were created to visualize associations. All statistical analyses were performed using R 4.0.2 (Vienna, Austria), all tests were two-sided and a P-value <0.05 was considered statistically significant.

## Supporting information

All Supplemental Tables and Figures

## Data Availability

All data and analyses are contained within this manuscript.

## Acknowledgements

We would like to thank all of our UNC CP donors, the staff at the UNC Blood Donation Center including Hannah Thaxton and Taylor A. Whitaker, and the many volunteers who contributed to this work. This project was supported by the UNC Health Foundation and the North Carolina Policy Collaboratory at the University of North Carolina at Chapel Hill with funding from the North Carolina Coronavirus Relief Fund established and appropriated by the North Carolina General Assembly. The NIH SeroNet Serocenter of Excellence Award, U54 CA260543, supported generation of laboratory data and the following investigators: AJM, LP, SN, SW, DMM, ADS, RB, and LAB. AJM was previously funded by an NIH NIAID T32 AI007151. SN is also funded by the following NIH grants: UNC Center for AIDS Research (P30 AI50410), NA-ACCORD COVID-19 Supplement (U01 AI069918). DRM is funded by an NIH F32 AI152296, a Burroughs Wellcome Fund Postdoctoral Enrichment Program Award, and was previously funded by an NIH NIAID T32 AI007151.

## Author Contributions

AJM performed experiments, data analysis and interpretation, and contributed to writing the manuscript. NG performed data and statistical analyses and contributed to writing the manuscript. DRM and XJH performed neutralization experiments and analysis. RR, PL, DRB, SDG and QG performed experiments. AL contributed to writing the manuscript. HR performed data analysis. CC and JS performed the N IgG Abbott assays. DMM and LB executed the clinical protocols, contributed to data analysis and interpretation, and contributed to the manuscript. JK, SW and YP executed the clinical protocols and coordinated donations and collection. SN contributed to statistical analyses and editing the manuscript. DVD edited the manuscript. CJP and TW generated, executed and completed data analysis of the PSV neutralization assay. Funding for the project was obtained by RB, ADS, DMM, and LAB.

