## Supplementary material for "Sex disparities and neutralizing antibody durability to SARS-CoV-2 infection in convalescent individuals": All Supplemental Tables and Figures

**Supplementary Information:**

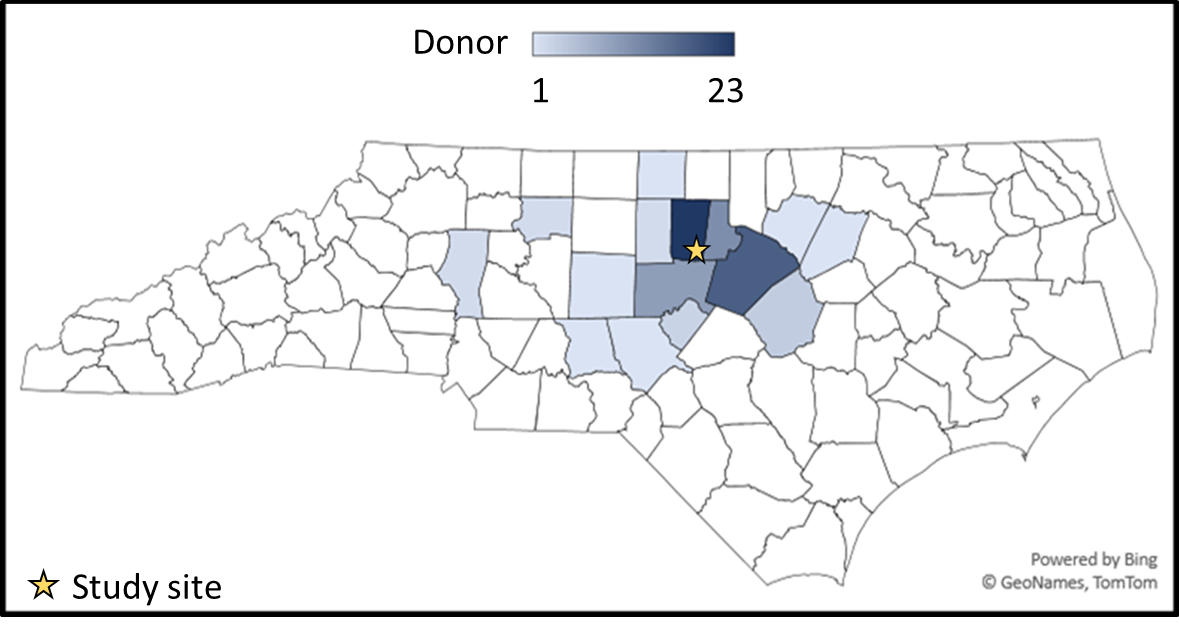

**Fig. S1 Map of donor zip codes.** A county map was generated using zip code data of donors that were located in North Carolina (n = 86), home site for study indicated by yellow star.

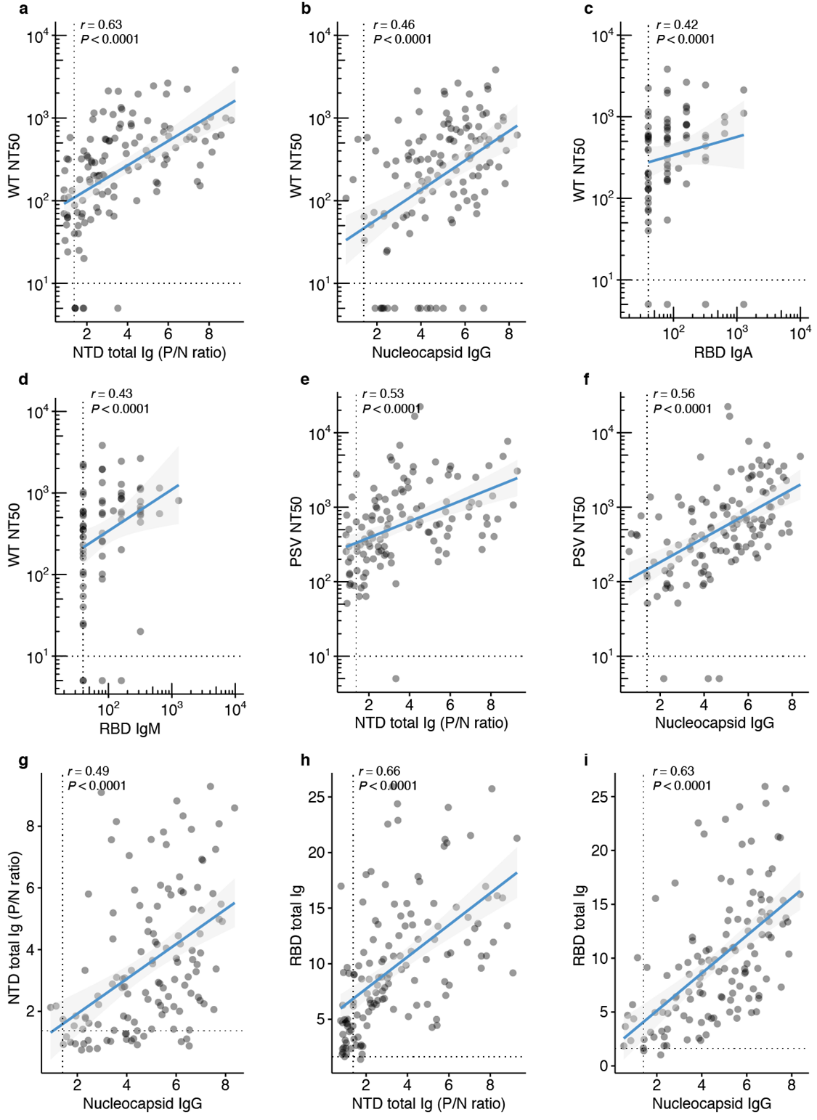

**Fig. S2 Correlation plots of antibody binding and functional assays**. **a-d**, Wild-type virus NT50 dilution plotted against antibody level. **e-f**, Pseudovirus NT50 dilution plotted against antibody level. **g**, NTD total Ig (P/N ratio) plotted against nucleocapsid IgG (Index value). **h,i**, RBD total Ig antibody level (end-point titer) plotted against antibody level. For **a-i**, Spearman’s rank correlation was used to calculate correlation coefficients (r) and P values (p), titers below LOD set to 5, all double-negative values removed, blue lines represent linear regression with 95% confidence interval (gray shading).

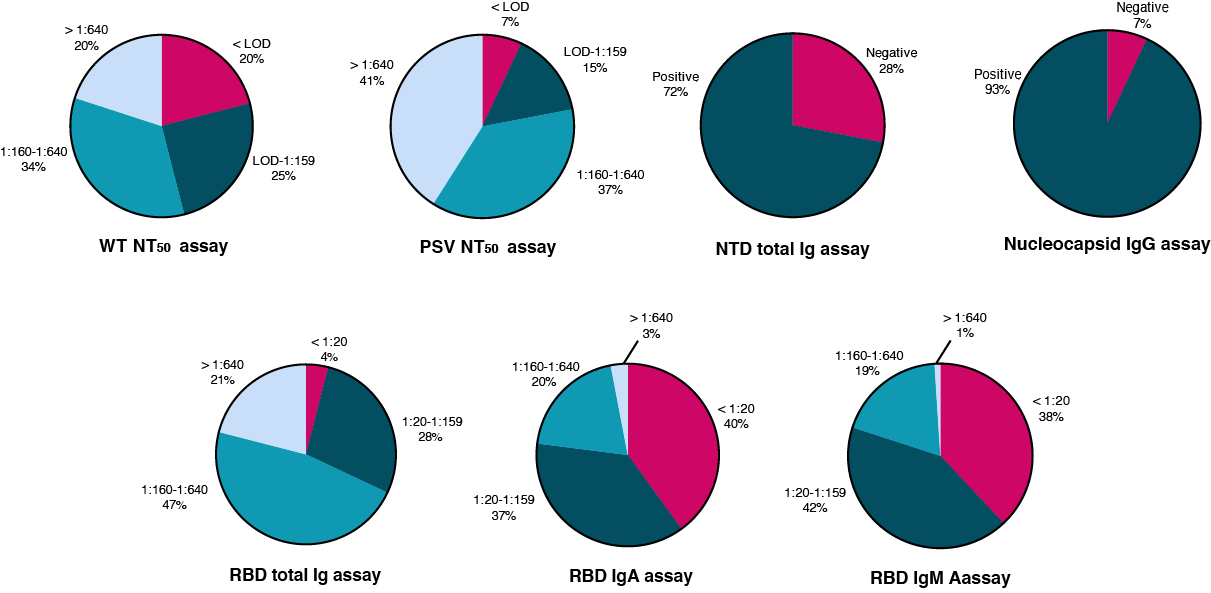

**Fig. S3 Neutralizing and binding antibody results broken down by assay and titer.**

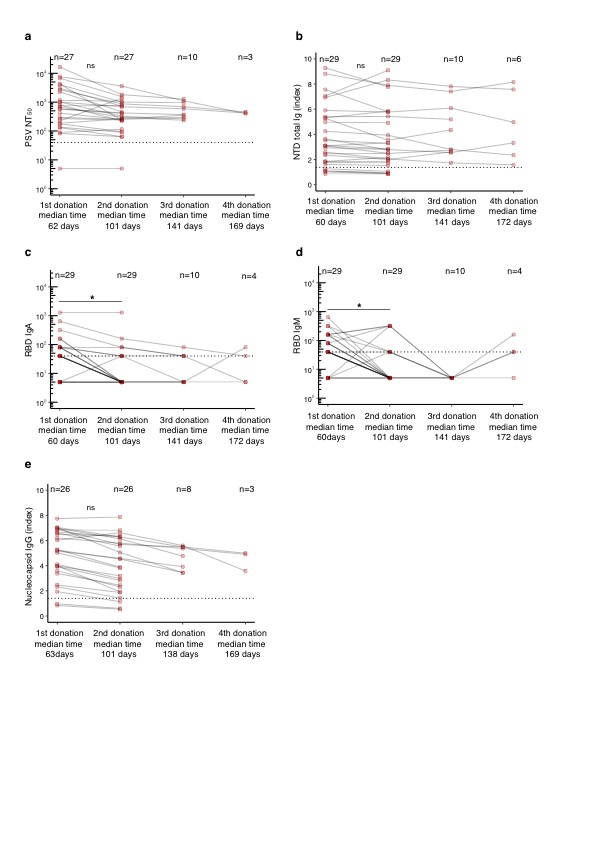

**Fig. S4 Antibody levels in sequential donors. a,** Pseudovirus NT50 dilution of sequential donors over four donations. **b**, NTD total Ig (P/N ratio) of sequential donors over four donations. **c**, RBD IgA (end-point titer) of sequential donors over four donations **d**, RBD IgM (end-point titer) of sequential donors over four donations **e**, Nucleocapsid IgG (Index value) of sequential donors over four donations. Titers are presented as geometric mean with geometric coefficient of variation. Statistical significance was determined using Mann-Whitney U-tests comparing donation 1 against donation 2 when matching donor data was available.

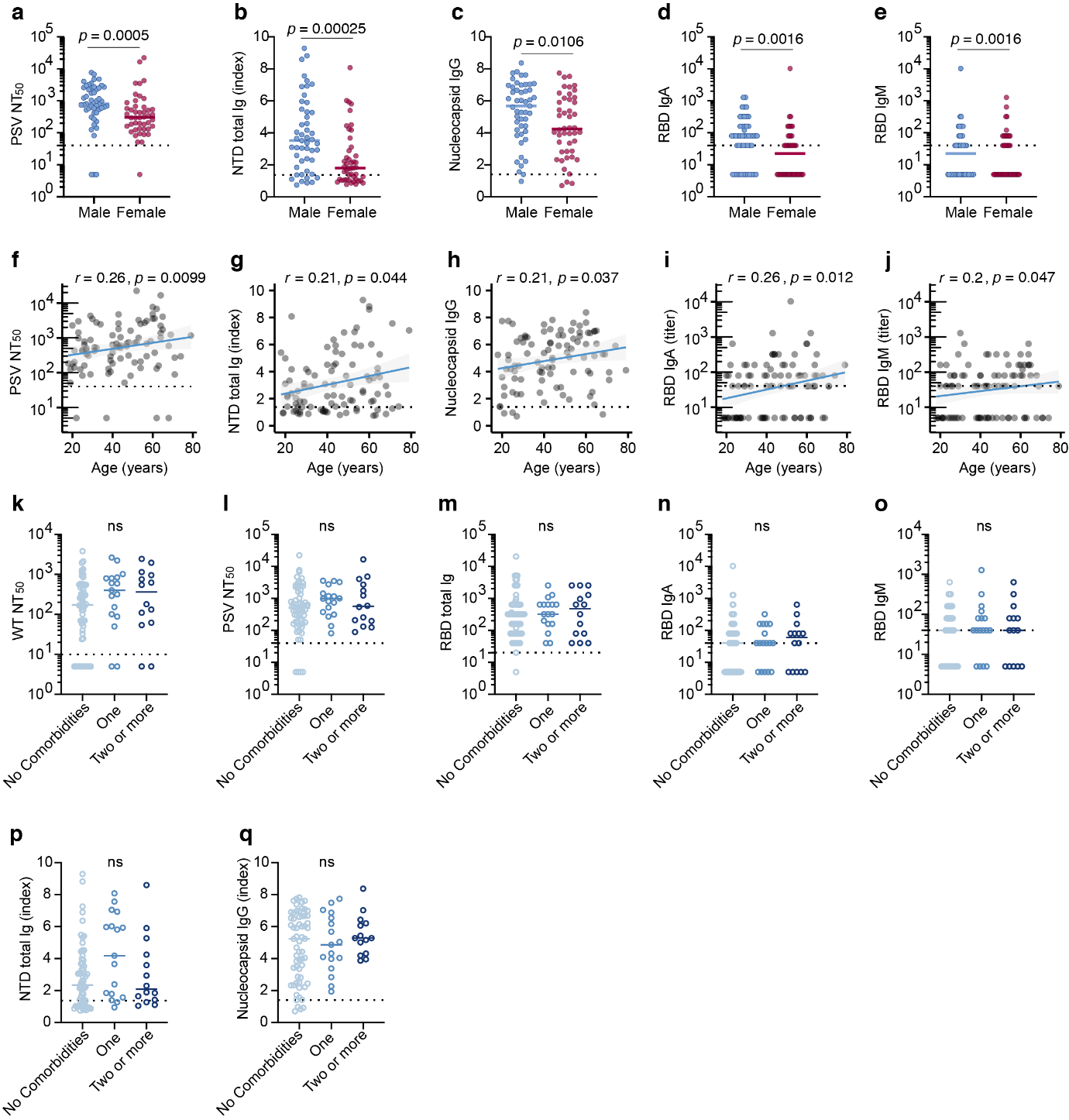

**Fig. S5 Additional demographic correlates of neutralization.** **a-e,** binding and neutralizing antibodies in males and females at first donation, **f-j,** Spearman correlation between age and binding and neutralizing antibodies at first donation, **k-q,** binding and neutralizing antibodies in donors with no reported comorbidities, one comorbid condition and two or more comorbidities. For **a-e** and **k-q** horizontal bars indicate median values. Statistical significance was determined using Mann-Whitney U-tests. For **f-j** spearman’s rank correlation was used to calculate correlation coefficients (r) and P values (p), blue lines represent linear regression fit with 95% confidence interval (gray shading).

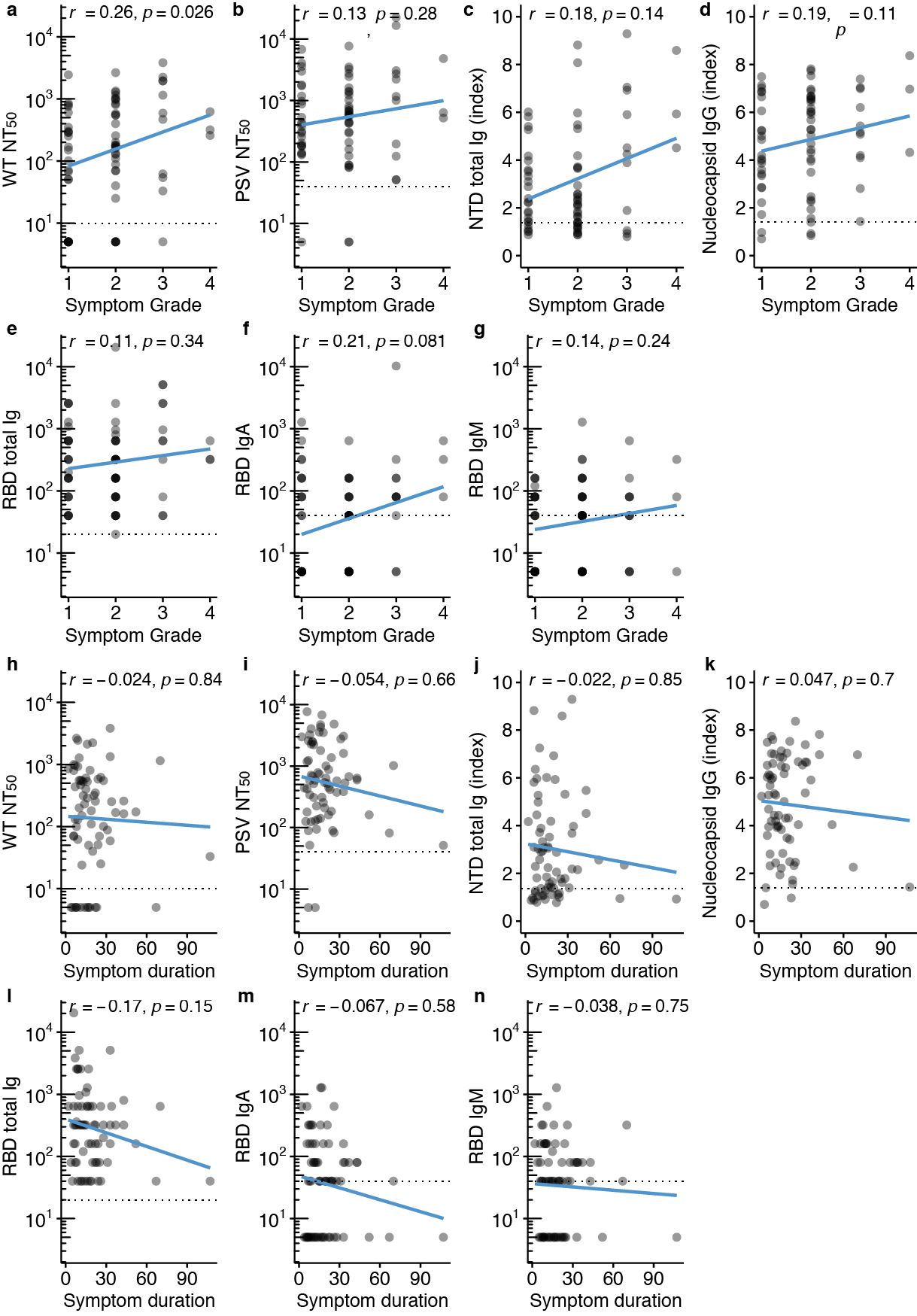

**Fig. S6 Symptom correlates of binding and neutralizing antibodies. a-g,** Spearman’s correlation between symptom grade and neutralizing or binding antibody level at first donation**, h-n,** Spearman’s correlation between symptom duration and neutralizing or binding antibody level at first donation. Spearman’s rank correlation was used to calculate correlation coefficients (r) and P values (*p*).

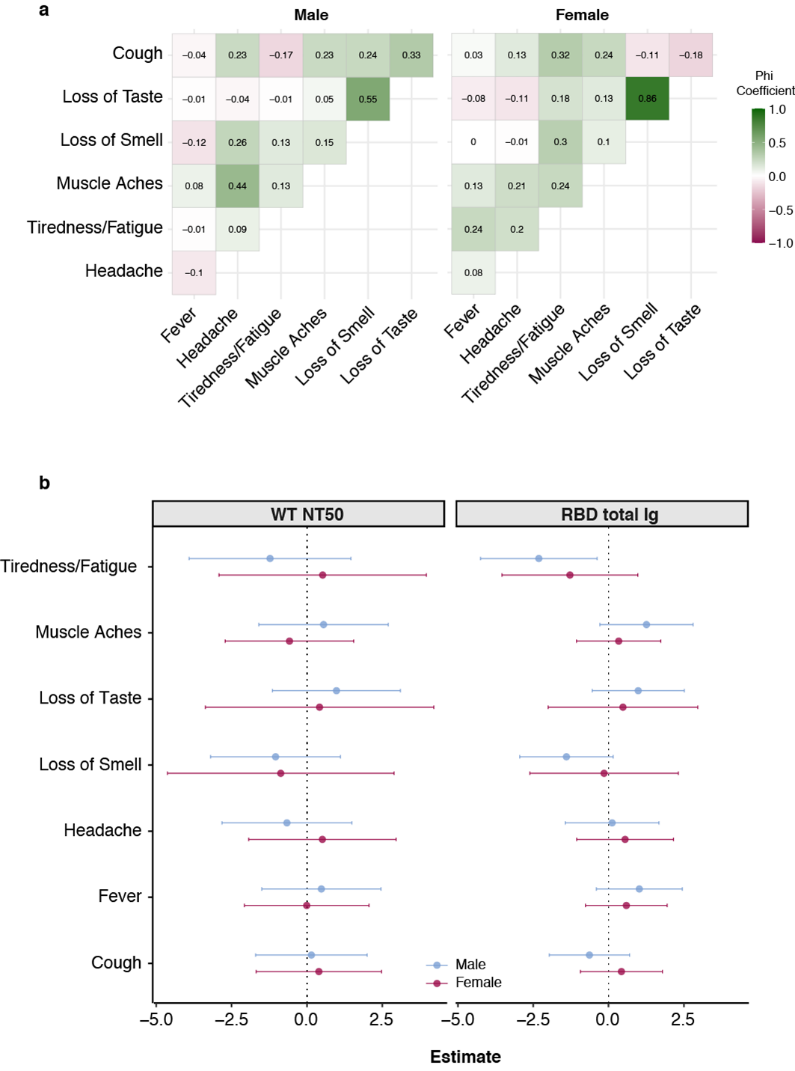

**Fig. S7 Symptomatology between sexes. a**, Heat map of phi coefficients examining the inter-relationship among reported symptoms in males (n=46) and females (n=44). Green colour indicates positive association and pink colour indicates negative association **b**, Forest plot showing the effect of reported symptoms in the levels of wild-type virus (left panel) and RBD total Ig (right panel) in males and females. Effects were calculated using linear regression models adjusted for age and time from symptom onset or PCR diagnosis.

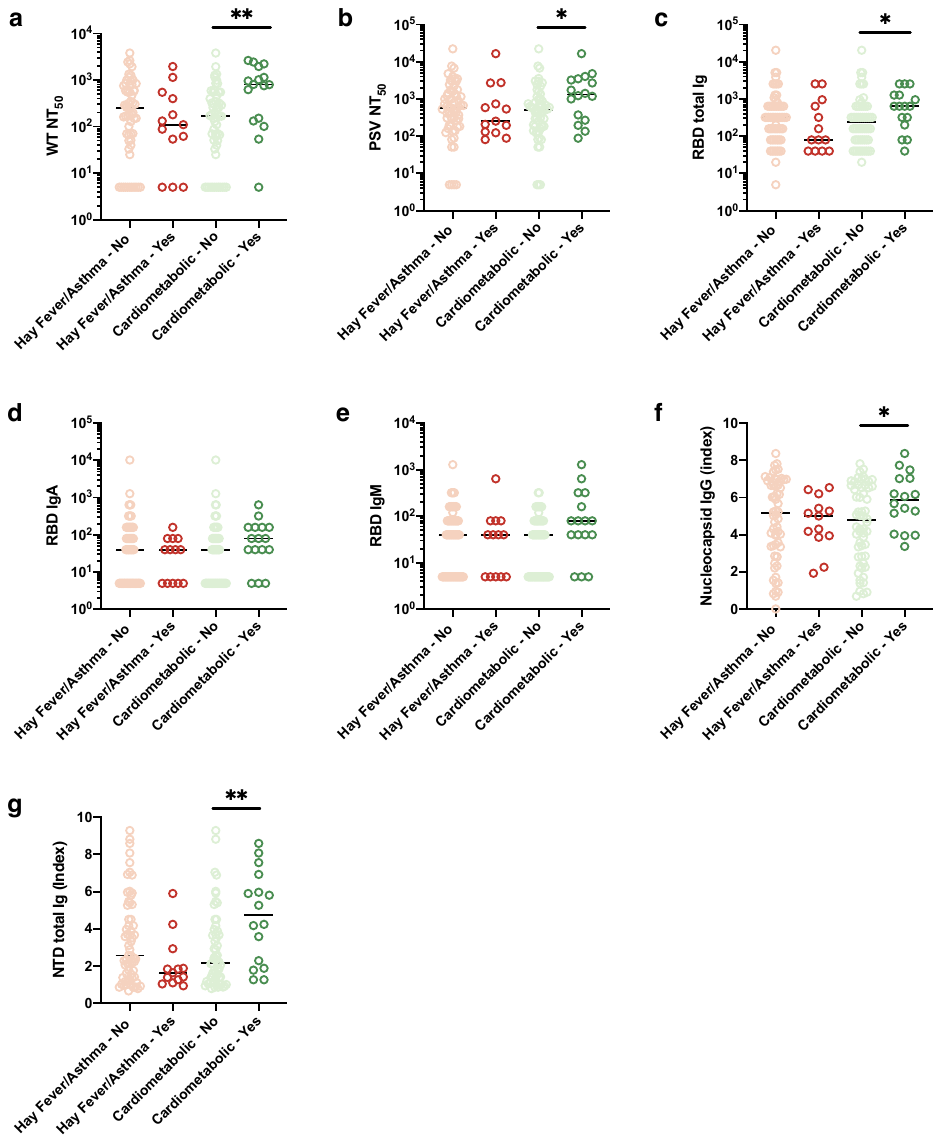

**Fig. S8 Antibody levels in donors with reported comorbid conditions** Donors were categorized into individuals with or without hay fever and/or asthma and into individuals with or without cardio-metabolic conditions (diabetes, obesity, hypertension, cardiovascular disease). Horizontal bars indicate median values. Statistical significance was determined within the two groups of conditions using Mann-Whitney U-tests.

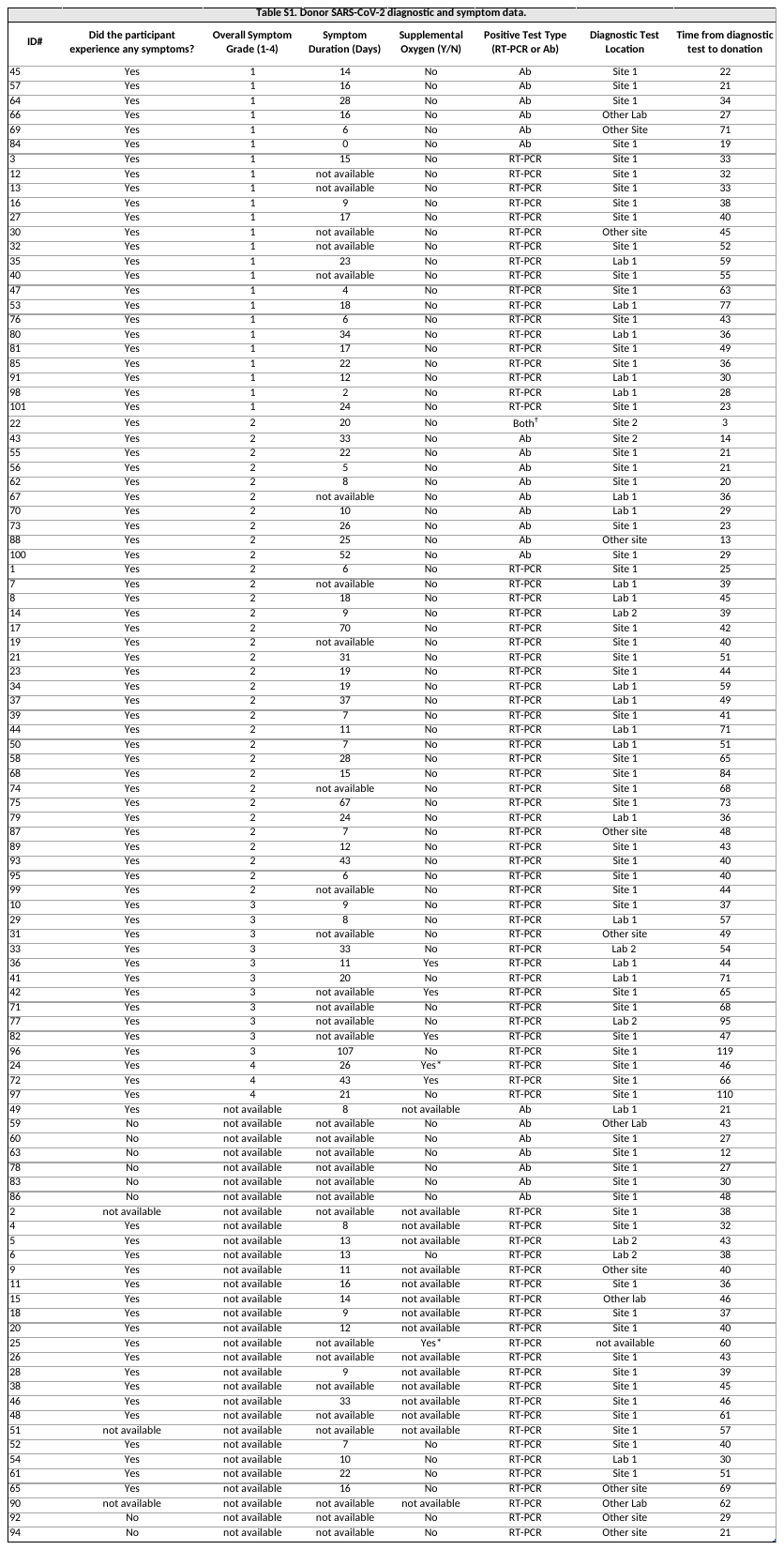

**Supplementary Table 1. Donor SARS-CoV-2 diagnostic and symptom data.** Site 1 indicates home site for this study. Lab 1 and 2 are testing labs located within North Carolina that accounted for at least five samples. Other site and other lab represent other US hospitals or laboratories. RT-PCR; Reverse-transcriptase polymerase chain reaction, Ab; Antibody.

*required mechanical ventilation

Ϯcould not provide RT-PCR test documentation

| **Table S2. Donor Comorbidities** | |
| --- | --- |
| Hay fever | 12 |
| High blood pressure | 10 |
| Asthma | 7 |
| Cancer (ever) | 7 |
| Diabetes | 6 |
| Obesity | 4 |
| Heart or cardiovascular disease | 3 |
| Inflammatory bowel disease | 3 |
| Blood disorder (like Sickle Cell) | 1 |
| Asplenia or hyposplenism | 1 |
| Liver disease | 1 |
| Cystic fibrosis | 0 |
| Kidney disease | 0 |
| Lupus, systemic lupus erythematosus | 0 |
| Multiple sclerosis | 0 |

**Supplementary Table 2. Donor comorbidity data.** Conditions reported by thirty four donors.

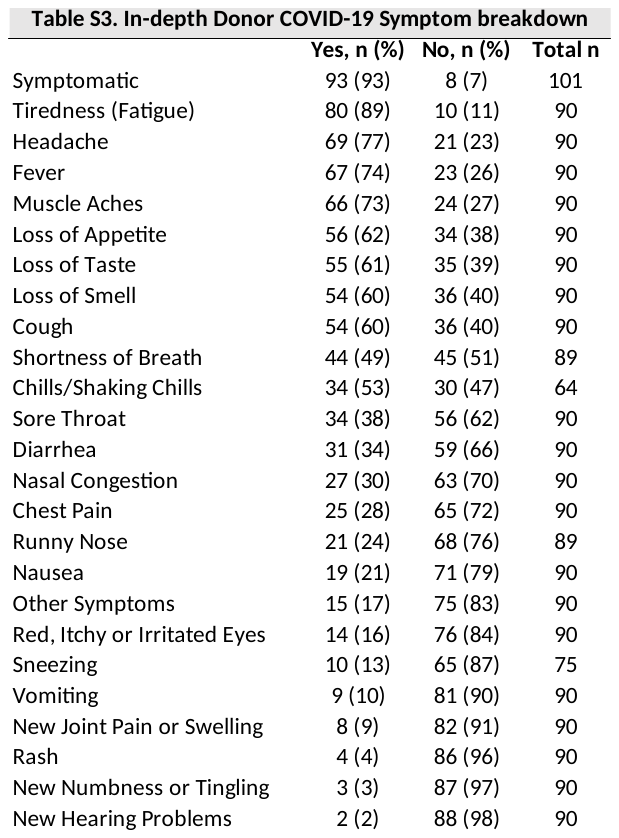

**Supplementary Table 3. In-depth donor COVID-19 symptom breakdown. “**Other symptoms” include: Abdominal pain (1), abdominal bloating (2), weight loss (1), leg pain, worsening vision (2), pruritic scalp (1), general symptom recurrence (1), seizures (1), excess thirst (1), urinary symptoms (1), syncopal episode (1), and severe nightmares (1). Numbers in parenthesis = n participants.

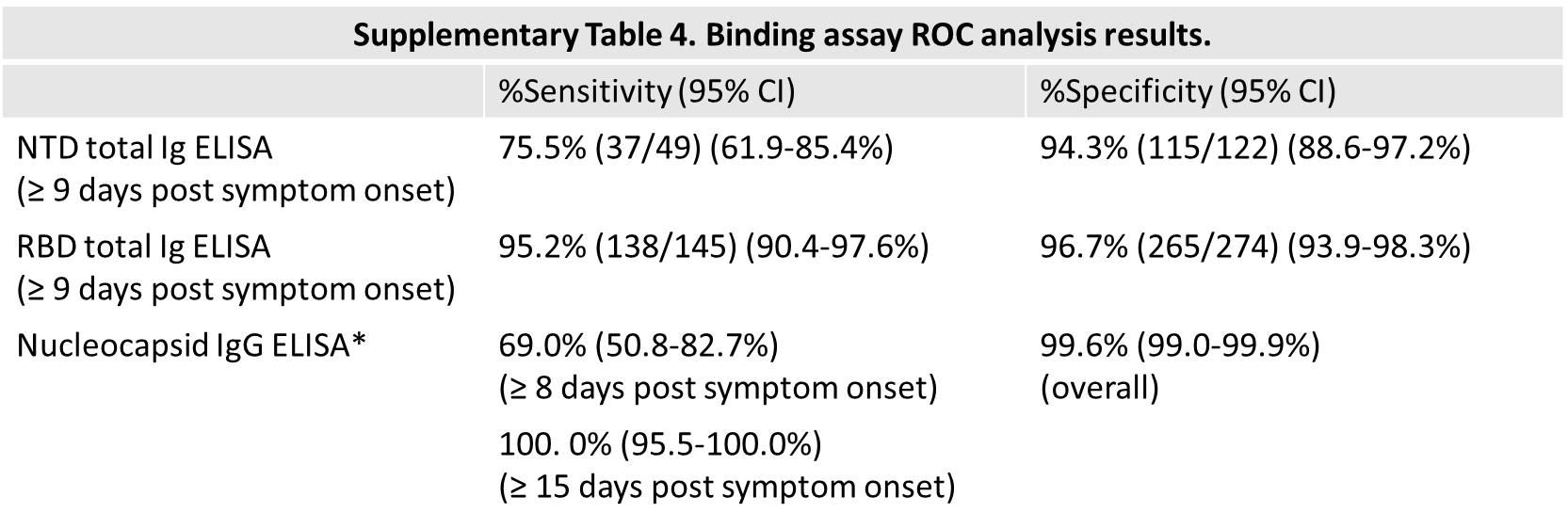

**Supplementary Table 4. Binding assay ROC analysis results.**

*Nucleocapsid data per previous publications and manufacturer (1-3).

1.	Nicol T*, et al.* (2020) Assessment of SARS-CoV-2 serological tests for the diagnosis of COVID-19 through the evaluation of three immunoassays: Two automated immunoassays (Euroimmun and Abbott) and one rapid lateral flow immunoassay (NG Biotech). *J Clin Virol* 129:104511.

2.	FDA (2020) EUA Authorized Serology Test Performance.

3.	Barzin A*, et al.* (2020) SARS-CoV-2 Seroprevalence among a Southern U.S. Population Indicates Limited Asymptomatic Spread under Physical Distancing Measures. *mBio* 11(5).
